# The effectiveness of isoniazid preventive treatment against tuberculosis among contacts of multidrug-resistant tuberculosis: A systematic review and individual-participant meta-analysis

**DOI:** 10.1101/2024.11.21.24317060

**Authors:** Leonardo Martinez, Neus Altet, Fadila Boulahbal, Joan A Cayla, Tsira Chakhaia, Pei-Chun Chan, Cheng Chen, Chi-Tai Fang, Greg Fox, Louis Grandjean, Djohar Hannoun, Anneke Hesseling, C. Robert Horsburgh, Li-Min Huang, Qiao Liu, Rufaida Mazahir, Chih-Hsin Lee, Li-Na Lee, Lisa Trieu, James A Seddon

**Affiliations:** Department of Epidemiology, School of Public Health, Boston University, Boston, Massachusetts, United States; Instituto de Investigación en Atención Primaria Jordi Gol, Barcelona, Spain; Groupe de recherche sur la tuberculose latente, Laboratoire National de Référence pour la Tuberculose, Institut Pasteur d’Algérie, Algiers, Algeria; Barcelona Tuberculosis Research Unit Foundation, Barcelona, Spain; National Center for Tuberculosis and Lung Diseases, Tbilisi, Georgia; Division of Chronic Infectious Disease, Taiwan Centers for Disease Control, Taipei, Taiwan; Department of Pediatrics, National Taiwan University Hospital, National Taiwan University College of Medicine, Taipei, Taiwan; Institute of Epidemiology and Preventive Medicine, College of Public Health, National Taiwan University, Taipei, Taiwan; Department of Chronic Communicable Disease, Center for Disease Control and Prevention of Jiangsu Province, Nanjing, Jiangsu Province, China; Department of Internal Medicine, National Taiwan University Hospital, Taipei, Taiwan; Sydney Medical School, Camperdown, NSW, Australia; Department of Infection, Inflammation and Immunity, Institute of Child Health, University College London, London, United Kingdom; Department of Information, National Institute of Public Health, Algiers, Algeria; Desmond Tutu TB Centre, Department of Paediatrics and Child Health, Stellenbosch University, Stellenbosch, South Africa; Department of Pediatrics, Jawaharlal Nehru Medical College, Aligarh Muslim University, Aligarh, India; Pulmonary Research Center, Wan Fang Hospital, Taipei Medical University, Taipei, Taiwan; Department of Internal Medicine, School of Medicine, College of Medicine, Taipei Medical University, Taipei, Taiwan; Department of Laboratory Medicine, Fu Jen Catholic University Hospital, Fu Jen Catholic University, New Taipei City, Taiwan; Division of Disease Control, New York City Department of Health and Mental Hygiene, Queens, NY, United States; Department of Infectious Disease, Imperial College London, United Kingdom

## Abstract

**Objective:** To evaluate the impact of isoniazid on incident tuberculosis in household contacts of MDR tuberculosis cases.

**Design:** Systematic review and individual-participant meta-analysis.

**Data sources:** MEDLINE, Web of Science, BIOSIS, and Embase without language restrictions for case-contact cohort studies of tuberculosis contacts.

**Eligibility criteria and data analysis:** Household contact tracing studies that investigated the development of tuberculosis in persons closely exposed to individuals with tuberculosis and followed for incident disease. Both retrospective and prospective cohort studies were included. Participants were included if they were exposed to someone with multidrug-resistant tuberculosis and were given either 6 months of isoniazid TPT or no TPT. Two reviewers independently assessed quality using a modified quality assessment of tool. We derived adjusted hazard ratios (aHRs) for incident tuberculosis using mixed-effects, multivariable Cox regression models with study-level random effects. The effectiveness of isoniazid TPT against incident tuberculosis was estimated through propensity score matching. We stratified our results by contact age, HIV, and *Mycobacterium tuberculosis* infection status.

**Main outcome measures:** Our primary outcome was incident tuberculosis in contacts exposed to tuberculosis (defined as a diagnosis >90 days after baseline). We derived adjusted hazard ratios (aHRs) for incident tuberculosis using mixed-effects, multivariable Cox regression models with study-level random effects.

**Results:** We included participant-level data from 4,945 contacts exposed to multidrug-resistant tuberculosis from eight countries. The effectiveness of 6 months of isoniazid TPT against tuberculosis in contacts of multidrug-resistant tuberculosis was 70% (aHR, 0.30; 95% CI, 0.16–0.56) and did not appreciably change with adjustment for additional potential confounders. Effectiveness was higher among contacts <18 years of age (aHR, 0.39; 95% CI, 0.18–0.87) compared to adult contacts (aHR, 0.44; 95% CI, 0.14–1.41). Effectiveness was 93% (aHR, 0.07; 95% CI, 0.02–0.52) in the first year of follow-up; effectiveness dropped to 80% (aHR, 0.20; 95% CI, 0.05–0.89) in the second year and was non-significant after two years (26% effectiveness; aHR, 0.74; 95% CI, 0.34–1.59).

**Conclusions:** Among almost 5,000 contacts of multidrug-resistant tuberculosis cases, isoniazid IPT was 70% effective against incident tuberculosis. Protection waned after 2 years of follow-up. These results have important implications for the clinical management of individuals exposed to multidrug-resistant tuberculosis and future clinical trials.

## Introduction

Following exposure to an individual with infectious tuberculosis, some contacts will develop *Mycobacterium tuberculosis* (*Mtb*) infection.^1,2^ *Mtb* infection is defined as having immunological sensitization to *Mtb* as measured by an interferon gamma release assay (IGRA) or tuberculin skin test (TST).^3^ Following infection, some individuals will progress to tuberculosis, characterized by symptoms and signs consistent with tuberculosis, frequently accompanied by radiological features and microbiological confirmation of the presence of *Mtb*. For over fifty years, drug treatment has been provided to individuals either with confirmed or suspected *Mtb* infection to prevent progression to disease, known as tuberculosis preventive treatment (TPT).^5,6^ There is a substantial evidence base for the efficacy and safety of isoniazid, rifamycins or a combination of isoniazid and a rifamycin to prevent disease progression in those exposed to drug-susceptible tuberculosis.^6,7^ The World Health Organization and most national tuberculosis guidelines advocate for the provision of TPT to contacts of tuberculosis who are judged to be at high risk of disease progression.^8^ This includes young children (<5 years), individuals living with HIV and those felt to be recently infected, where the risk of disease progression is highest. However, management of individuals exposed to drug-resistant tuberculosis is unclear.

Multidrug-resistant (MDR) tuberculosis is defined as disease caused by *Mtb* resistant to at least isoniazid and rifampicin and each year an estimated 500,000 individuals develop MDR tuberculosis, with several million of their close contacts subsequently exposed to, and infected with, MDR-*Mtb*.^9,10^ As MDR tuberculosis is, by definition, resistant to the two main drug classes used for drug-susceptible TPT, it is unclear what drug should be given to prevent tuberculosis progression in exposed persons. International guidance is mixed with the World Health Organization currently suggesting that MDR-TPT can be considered in high-risk MDR tuberculosis contacts following an individual patient risk assessment.^8,11^ Two clinical trials have recently been completed that have evaluated whether daily levofloxacin given for six months prevents disease progression in household contacts of MDR tuberculosis. V-QUIN recruited mainly IGRA-positive adults exposed in their household to MDR tuberculosis while TB-CHAMP took place in South Africa and recruited mainly child contacts <5 years, irrespective of IGRA status.^12–15^ Both trials demonstrated a substantial effect size of the intervention, yet neither trial reached a threshold of precision that met statistical significance.^14,15^ Planned meta-analysis of the two trials demonstrated significant efficacy of levofloxacin. Despite these promising results, there are challenges to the use of this drug for this indication.

Unlike isoniazid and the rifamycins, levofloxacin is a broad-spectrum antibiotic that has efficacy against a wide range of Gram-positive and Gram-negative bacteria, and long-term use has the potential to substantially disrupt the gut and respiratory microbiome with unknown implications. In addition, there is potential to drive resistance to the fluoroquinolone class of antibiotics in non-tuberculosis bacteria which could make treatment of serious, life-threatening infections more challenging. Levofloxacin is not currently available in many low resource settings and, in tuberculosis programmes, is generally reserved for the treatment of MDR tuberculosis. For all of these reasons, it would be important to explore alternative drug options in preventing tuberculosis in MDR tuberculosis contacts.

Recent empirical research has suggested that isoniazid may lead to a risk reduction of incident tuberculosis among close tuberculosis contacts.^16^ To evaluate the impact of isoniazid on incident tuberculosis in household contacts of MDR tuberculosis cases, we carried out an analysis within a large individual participant dataset of individuals with close exposure to persons with multidrug-resistant tuberculosis.

## Methods

### Search strategy and selection criteria

In this project, we used the same data collection and data collation methods as previously published systematic reviews and individual participant meta-analyses from this consortium investigating the development of tuberculosis among persons closely exposed to tuberculosis.^17–19^ Briefly, this systematic review and individual participant data meta-analysis follows Preferred Reporting Items for Systematic Reviews and Meta-Analyses guidelines for individual patient data meta-analyses. We searched MEDLINE, Web of Science, BIOSIS, and Embase without language restrictions for case-contact cohort studies of tuberculosis contacts published between Jan 1, 1998, and April 7, 2018. The 20-year timeframe was chosen on the basis of the expected availability of individual participant data. To prioritize an incident tuberculosis outcome, we restricted our search to cohort studies only. Both case-control studies and outbreak reports were excluded. Search terms included “*Mycobacterium tuberculosis*”, “TB”, “tuberculosis”, and “contact” and can be found in full in the appendix. Partnered with our systematic review, we also reviewed other systematic reviews and review articles of tuberculosis contact investigations. We inspected their reference lists for eligible articles. We included data that were unpublished (found through discussions with authors and data experts, with data located in data storage repositories, conference abstracts, and dissertations) if eligible.

Two independent reviewers assessed the quality of each study using a modified rubric of the Newcastle-Ottawa scale (Appendix). Each study was judged on the basis of a 9-point scale using three broad criteria: selection of participants (4 points), comparability of studies (2 points), and ascertainment of outcome of interest (3 points). High study quality was defined as a score of 6 or greater, moderate quality as 3 to 6 points, and low quality as below 3 points. Discrepancies were resolved by re-evaluating the study for consensus. This study follows PRISMA-IPD guidelines for individual-participant data reporting. The study protocol is registered with PROSPERO (CRD42018087022).

### Study definitions

Participants were characterized as being exposed to tuberculosis if they were reported to be a close contact (either living in the same household or having substantial interaction outside the household) of a person with microbiologically or radiologically diagnosed pulmonary tuberculosis. Investigators from each study defined exposure and index case diagnoses; we used the study definitions assigned to each cohort. We used each study’s classification of tuberculosis. Prevalent tuberculosis was defined as any diagnosis of tuberculosis at the initial visit or within 90 days of baseline evaluation. Incident tuberculosis was defined as a new tuberculosis case diagnosed more than 90 days after the initial evaluation. We restricted to only contacts of MDR tuberculosis and only included TPT regimens using isoniazid for six months. Tuberculosis infection was defined by a positive QuantiFERON-TB Gold In-Tube test (IFNγ nil value ≥0.35 IU/mL), T-SPOT.TB test (nil spots minus antigen spots ≥8), or tuberculin skin test (≥10 mm induration). Countries were classified into income levels by use of World Bank 2020 definitions (high-income, upper-middle-income, lower-middle-income, and low-income countries). MDR tuberculosis was defined as resistant to, at a minimum, rifampin and isoniazid, the two most potent tuberculosis-related drugs.

### Data analysis

Individual participant data for a prespecified list of variables, including the characteristics of the exposed contact, the index case, and the environment, were requested from authors of all eligible studies. We pooled individual participant-level data from all included cohorts. The primary aim of our analysis was to estimate the overall and adjusted effectiveness of six months of isoniazid TPT in preventing incident tuberculosis in MDR tuberculosis contacts. Therefore, our primary outcome was incident tuberculosis diagnosed more than 90 days from baseline. All participants developing tuberculosis before this time point (including at baseline) were excluded. We calculated follow-up time from the first baseline visit to development of tuberculosis, loss to follow-up, death, or study completion

For the primary analyses, we derived adjusted hazard ratios (aHRs) using mixed-effects, binary, multivariable Cox regression and parametric survival-time models incorporating study-level random effects into each model. We assumed a conditional Bernoulli distribution of the response given the random effects. We conducted a propensity score analysis when evaluating the protective effect of TPT. In this analysis matching was based on individual-level covariates of contact age, contact sex, contact previous tuberculosis, and whether data were collected prospectively or retrospectively. We then matched contacts who began isoniazid TPT with individuals who did not receive any TPT, using a nearest neighbor matching algorithm. In this matched cohort, we estimated covariate-adjusted risk of incident tuberculosis between groups when examining the protective effectiveness of isoniazid TPT. We conducted secondary multivariable regression analyses into key subgroups to assess whether effectiveness was modified by risk profile. We further stratified our analysis by contact age, *M tuberculosis* infection status, contact prior tuberculosis, World Health Organization region, and background tuberculosis incidence levels.

To assess the durability of the treatment effect, follow-up time was split into three time groups (<12 months, 12–23 months, and 24 months and after). We specified the effects of isoniazid by follow-up time and derived adjusted hazard ratios (aHRs) for each time interval.

## Results

### Systematic Review

In total, 14,927 original study reports from our database searches were identified (**Figure 1**). We screened 9,753 through an exclusionary keyword algorithm. We tested the exclusionary words approach for accuracy by implementing it on a random list of 100 titles that were also manually screened for eligibility to our study. Our exclusionary algorithm eliminated all articles that were excluded by manual screening with 100% specificity. We reviewed 512 full text articles published on or after January 1, 1998. 80 study groups were contacted for individual participant data and study groups from 48 studies agreed to share their data, which were collated into a single database of 461,285 individuals exposed to tuberculosis. 36 studies were excluded because they were without data on the drug resistance status of the index case, had no index cases with drug resistance, or did not provide information on TPT to contacts of drug-resistant tuberculosis. In total, nine cohort studies from our larger meta-analysis were included in this meta-analysis.

**Figure 1.**
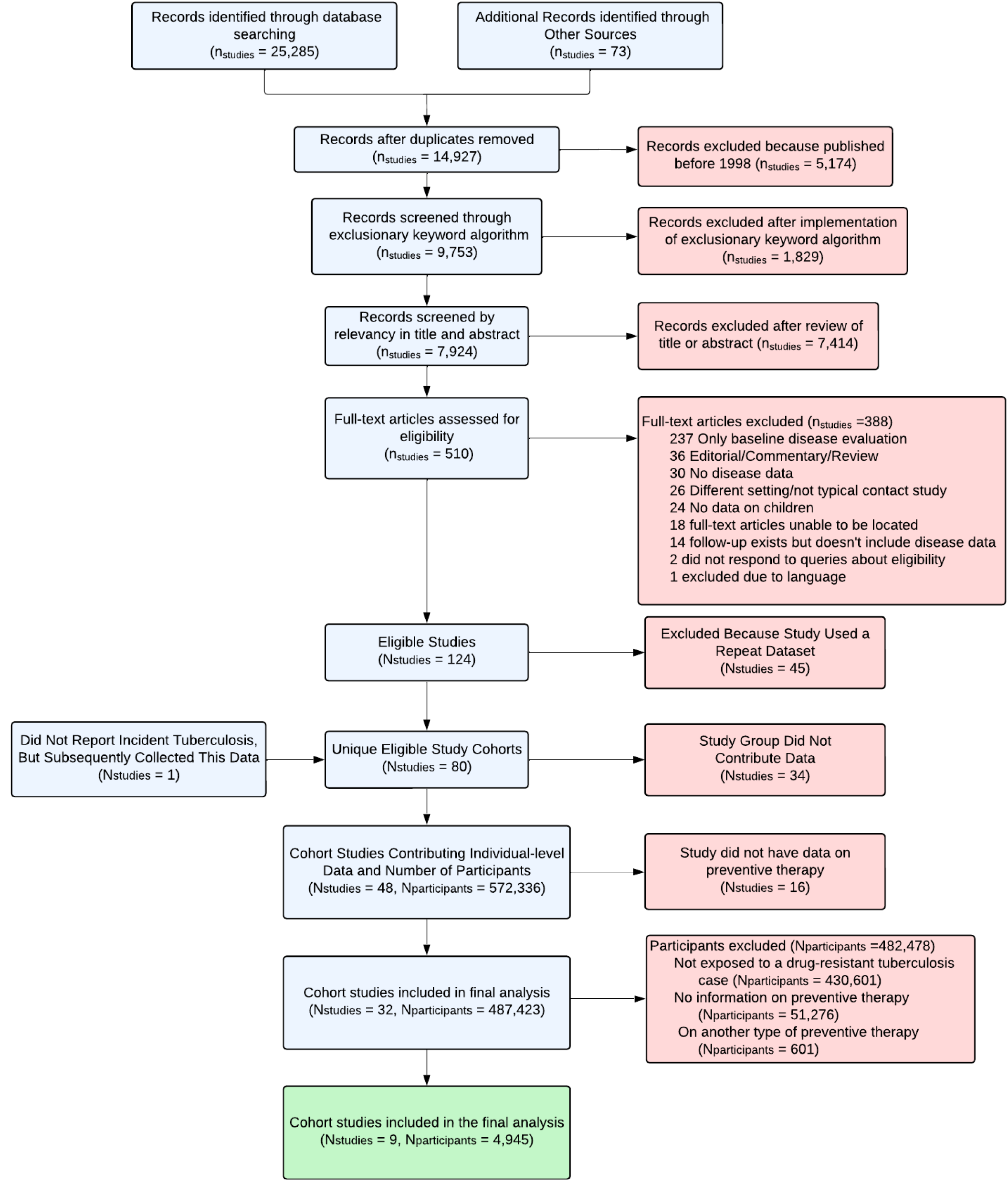
Flowchart of Systematic Search Process. Excluded full-text articles may have had more than one reason for exclusion, but only one reason for exclusion was listed for each excluded manuscript. Exclusionary keyword algorithm is described in the Supplementary Appendix.

Five (56%) of the nine cohorts used a prospective cohort study design. Three (33%) cohorts were in countries with a background burden of >100 incident cases per 100 thousand people. Seven (78%) of the nine cohorts performed IGRA or TST on contacts. Overall, study quality was considered for eight of nine studies (one study was a conference abstract and was not reviewable); all of these studies were considered at either a moderate (N_cohort_=3; 38%) or high (N_cohort_=5; 62%) study quality (Appendix).

In total, 4,945 participants were followed for incident tuberculosis for 12,316 person-years (**Table 1**). The median age of contact participants was 29 years (IQR, 14–47). Half of participants were 30 years and older (N=2,431; 50%). Among 4,341 contacts tested for HIV, 68 (5%) were positive. One-third of participants were evaluated with a test for *M. tuberculosis* infection (N=1,618; 33%). Most participants (N_participants_=3,186; 65%) were in settings with a background tuberculosis incidence >100 cases per 100 thousand persons. Among all participants 722 (15%) were given isoniazid TPT while 4,224 were not given TPT.

**Table 1.**
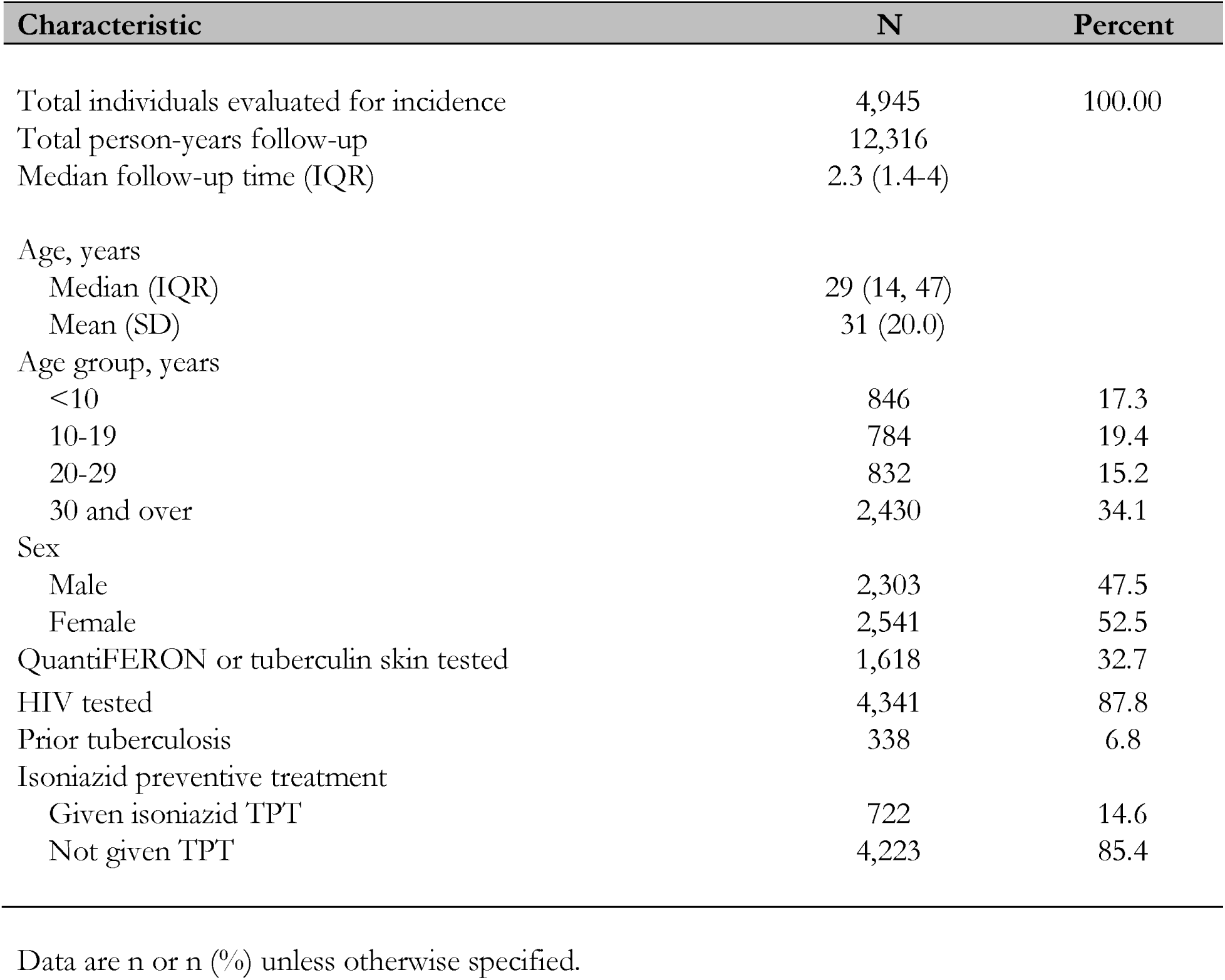
Demographic characteristics of included participants exposed to an individual with multidrug-resistant tuberculosis from included studies.

| Characteristic | N | Percent |
| --- | --- | --- |
| Total individuals evaluated for incidence | 4,945 | 100.00 |
| Total person-years follow-up | 12,316 |  |
| Median follow-up time (IQR) | 2.3 (1.4-4) |  |
| Age, years |  |  |
| Median (IQR) | 29 (14, 47) |  |
| Mean (SD) | 31 (20.0) |  |
| Age group, years |  |  |
| <10 | 846 | 17.3 |
| 10-19 | 784 | 19.4 |
| 20-29 | 832 | 15.2 |
| 30 and over | 2,430 | 34.1 |
| Sex |  |  |
| Male | 2,303 | 47.5 |
| Female | 2,541 | 52.5 |
| QuantiferON or tuberculin skin tested | 1,618 | 32.7 |
| HIV tested | 4,341 | 87.8 |
| Prior tuberculosis | 338 | 6.8 |
| Isoniazid preventive treatment |  |  |
| Given isoniazid TPT | 722 | 14.6 |
| Not given TPT | 4,223 | 85.4 |

Data are n or n (%) unless otherwise specified.

Over 12,316 person-years of follow-up, 151 persons developed tuberculosis (1.2 cases per 100 person-years). Among tuberculosis cases, 138 were not given TPT and 13 were given isoniazid TPT. Of these, 70/151 [46%] were diagnosed in the first year of follow-up. 35 cases developed tuberculosis in the second year while 46 cases developed disease subsequently after year 2 (**Figure 2**).

**Figure 2.**
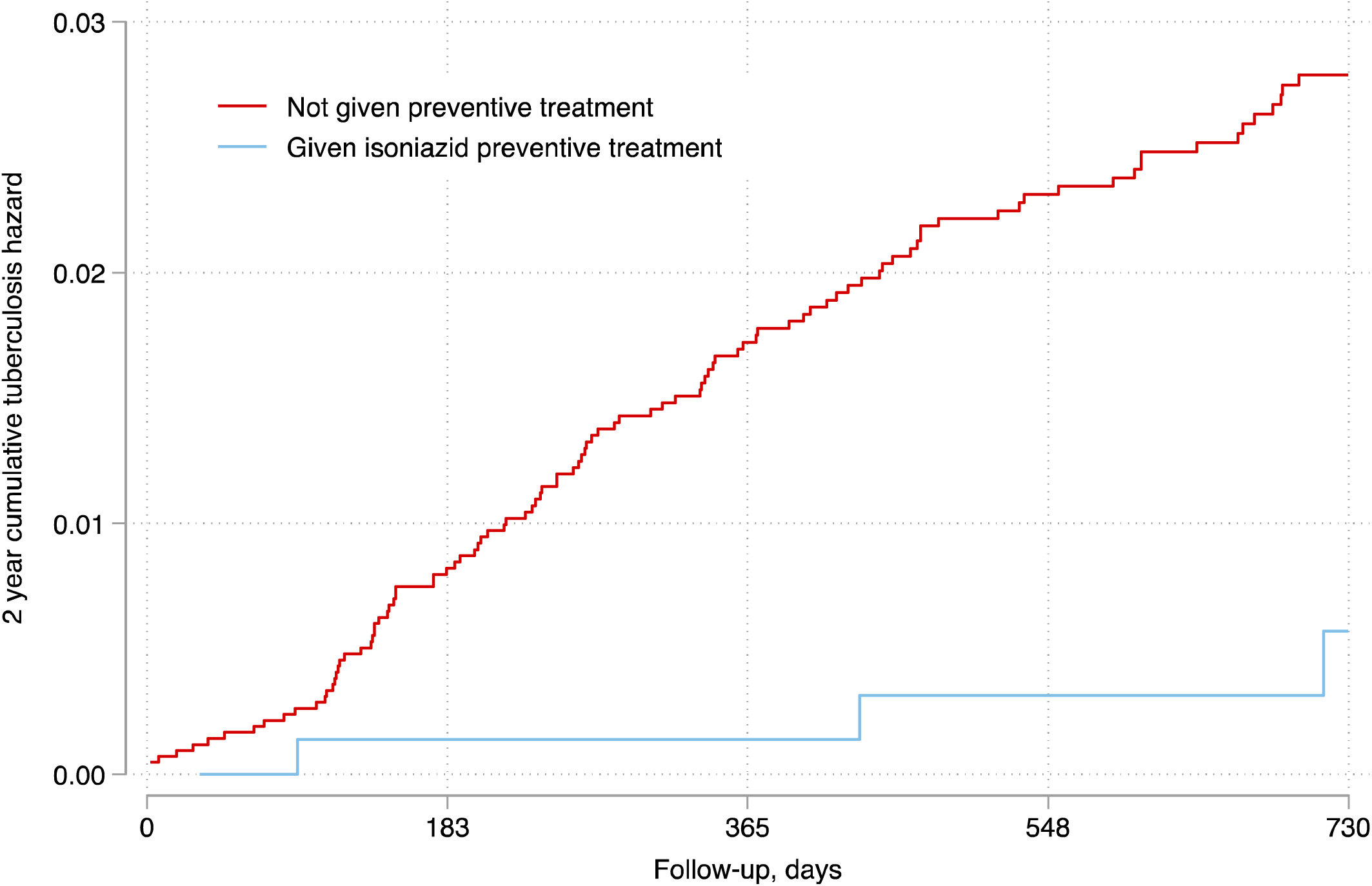
Cumulative hazard for isoniazid tuberculosis preventive treatment against incident tuberculosis among contacts of multidrug-resistant tuberculosis. The y-axis represents the cumulative hazard of tuberculosis. We restricted follow-up to the first 2 years after baseline. This graph represents a crude, non-adjusted Kaplan-Meier curve which does not take into account study-level random effects.

Among the 151 progressors, 67 (44%) had information on drug-susceptibility status. Among these, 65 were not given TPT; 49 (73%) of these progressors developed MDR tuberculosis while 7 (11%) developed drug-susceptible tuberculosis. The remaining cases were either mono-resistant (n=2; 4%) or polyresistant (n=7; 11%). Among the two progressors with drug-susceptibility status that were given TPT, both developed MDR tuberculosis.

In an age-adjusted mixed-effects model, the effectiveness of isoniazid TPT against tuberculosis in contacts of multidrug-resistant tuberculosis (N_participants_=4,945) was 68% (aHR, 0.32; 95% CI, 0.18-0.58) (**Table 2**). This protection did not appreciably change with adjustment for additional potential confounders. For example, when this model was additionally adjusted for sex, protection against incident tuberculosis was 70% (aHR, 0.44; 95% CI, 0.24–0.80). In a fully adjusted model, protection remained at 70% (aHR, 0.30; 95% CI, 0.16–0.55). The number needed to treat with isoniazid to prevent one incident case among contacts of MDR tuberculosis cases was 40.

**Table 2.** Effectiveness of isoniazid on risk of incident tuberculosis in household contacts of multidrug-resistant tuberculosis cases.

|  | **Total N** |  | **Person-years**<br>**of follow-up** | **N,**<br>**contacts** | **N, progressing**<br>**to disease** | **Age-adjusted HR**<br>**(95% CI)** | **Fully-adjusted HR**<br>**(95% CI)** |
| --- | --- | --- | --- | --- | --- | --- | --- |
| All participants | 4,945 | Not given<br>TPT | 10,653 | 4,223 | 140 | 1 (Referent) | 1 (Referent) |
|  |  | Given INH | 1,663 | 722 | 13 | 0.32 (0.18-0.58) | 0.35 (0.18-0.67) |
| Child <18 years | 1,467 | Not given<br>TPT | 1,928 | 884 | 22 | 1 (Referent) | 1 (Referent) |
|  |  | Given INH | 1,234 | 583 | 10 | 0.44 (0.20-0.93) | 0.39 (0.18-0.87) |

INH, Isoniazid; HR, hazard ratio; TPT, tuberculosis preventive treatment.

Effectiveness was similar among younger contacts <18 years of age (aHR, 0.39; 95% CI, 0.18-0.87) compared to adult contacts (aHR, 0.44; 95% CI, 0.14–1.41). Among 423 children <5 years old, none developed incident tuberculosis among the isoniazid TPT group (0/176; 0%) while five developed incident tuberculosis among children that were not given TPT (5/247; 2%). The effect of isoniazid TPT was highest in settings with a background tuberculosis incidence >100 cases per 100 thousand persons (risk difference, 2.7; 95% CI, 1.2– 4.1). In settings with a background tuberculosis incidence of <50 and 50–100 cases per 100 thousand persons, there were no incident cases among the group given isoniazid preventive treatment and therefore a measure of effect could not be calculated. Other covariates of interest did not modify the effectiveness of isoniazid TPT; however, in some of these subgroup analyses, there was a lack of statistical power (Appendix).

Although assessing an effect of isoniazid TPT was difficult to show in any one cohort due to statistical power, the effect was largely consistent across the largest cohorts. For example, in the three largest included cohorts, the increased risk difference of incident tuberculosis among contacts without TPT versus contacts taking isoniazid TPT was 3.7% (95% CI, 2.5–4.9), 2.9% (95% CI, 0.9–4.8), and 1.2% (95% CI, 0.5–1.9), respectively.

The effect of isoniazid on the risk of tuberculosis was greatest in the first year of follow-up (aHR 0.07; 95% CI, 0.02–0.52). The effect decreased over follow-up time (**Figure 3**). At 12–23 months of follow-up, effectiveness was 80% (aHR, 0.20; 95% CI, 0.05–0.89). After 24 months of follow-up, effectiveness was 26% but was no longer statistically significant (aHR, 0.74; 95% CI, 0.34–1.59).

**Figure 3.**
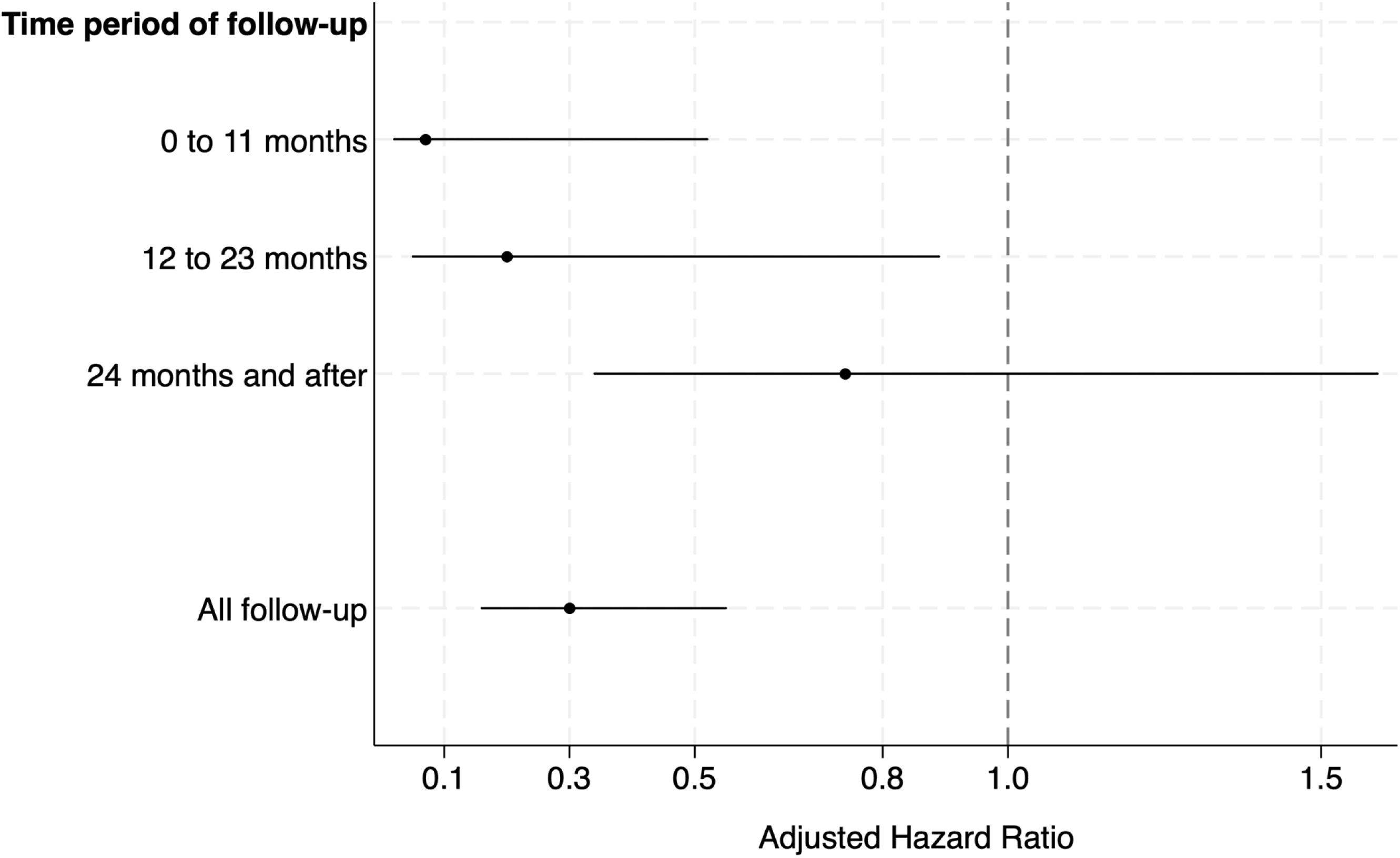
Effectiveness of isoniazid preventive treatment against incident tuberculosis among contacts exposed to multidrug-resistant tuberculosis, by period of follow-up. To assess the durability of the treatment effect, follow-up time was split into three time groups (<12 months, 12–23 months, and 24 months and after). We specified the effects of isoniazid by follow-up time and derived adjusted hazard ratios (aHRs) for each time interval. Models integrated a study-level random effect and adjusted for contact age, contact sex, and study design.

## Discussion

We analyzed the impact of isoniazid on the risk of incident tuberculosis in nearly 5,000 household contacts of individuals with MDR tuberculosis, assessed over 12,316 participant years of follow up. Our study demonstrated that isoniazid significantly reduced the risk of incident tuberculosis by 70% and that children gained a greater risk reduction from isoniazid than adults. This association was especially apparent among contacts in high tuberculosis burden settings. Importantly, the duration of protection persisted for up to two years after exposure.

We found that overall isoniazid was 70% effective in protecting against incident tuberculosis among contacts of MDR tuberculosis. These results are similar to a study carried out in Peru published in 2020, which recruited and followed up 652 children and adolescents (<20 years) exposed to MDR tuberculosis.^16^ This study found that isoniazid reduced the risk of incident tuberculosis in household contacts of individuals with MDR tuberculosis by 81% (HR, 0.19; 95% CI, 0.05-0.66). A previous study in Rio de Janeiro had retrospectively assessed 218 adult and child contacts of MDR tuberculosis, of whom 45 had been given isoniazid.^20^ This study failed to find any protective effect of isoniazid with two of 45 contacts who had been given isoniazid developing disease (1.2 per 1,000 person-years), compared to 13 of 145 who did not receive isoniazid progressing (1.7 per 1,000 person-years; p=0.47). There have been isolated examples of individuals exposed to MDR tuberculosis progressing to disease, despite preventive treatment with first-line drugs.^21^ Given the *in vitro* resistance implicit in the definition of MDR tuberculosis, most national and international guidelines have, for many years advised against isoniazid as TPT for individuals exposed to MDR tuberculosis.

With the data available, it is not possible to interrogate underlying mechanisms for the results seen in this study. However, several are plausible. First, in high tuberculosis incidence contexts, individuals are exposed to *Mtb* frequently and many will have been exposed to, and infected by, *Mtb* in the years before someone in their household has been diagnosed with MDR-TB. Given that drug-susceptible tuberculosis is more common than MDR tuberculosis in almost all contexts, it is likely that previous infection would have been with a drug-susceptible strain. Successful treatment of this drug-susceptible isolate with isoniazid would therefore reduce the incidence of incident tuberculosis. Our finding that effectiveness was highest in studies with a high background tuberculosis burden supports this hypothesis. However, the low sample sizes (and disease events) in settings with a lower burden of tuberculosis from our consortium makes it difficult to definitively conclude this mechanism. Further research is needed to understand whether isoniazid is effective against progression among contacts exposed to MDR in settings with a low-burden of tuberculosis. Second, isoniazid resistance is mediated through multiple resistance mechanisms, encoded for by several genetic mutations to the *Mtb* genome.^22,23^ Multiple distinct mutations in the *katG* gene and *inhA* promoter region lead to resistance but the resulting minimum inhibitory concentration can be highly variable depending on exact site of mutation and isoniazid can still be effective against some of these strains. Finally, the detection of *in vitro* resistance, either through genotypic or phenotypic evaluation, does not always translate into the drug being ineffective *in vivo* and novel mechanisms of action for isoniazid have been proposed in which the drug has efficacy not only by killing *Mtb*, but also by interacting with the host immune system to promote *Mtb* death.^24^

An increasing body of empirical data has demonstrated the efficacy and safety of alternative regimens as TPT for contacts of MDR tuberculosis. Recently completed trials – V-QUIN and TB-CHAMP – provide compelling evidence for fluoroquinolone TPT for MDR tuberculosis contacts.^14,15^ Despite this, there is still likely to be an important role for isoniazid in contexts where fluoroquinolones are unavailable, too expensive, or if tuberculosis programmes or, individual clinical staff, are unwilling to endorse usage. There may be some advantages to using isoniazid as MDR-TPT, given fewer non-specific effects on antimicrobial resistance and less disruption to the microbiome. Isoniazid may also be more available at lower levels of care and appropriate formulations may be stocked more regularly in remote health facilities. Having more than one option as TPT for contacts exposed to MDR tuberculosis is likely to be useful for most tuberculosis programmes and clinical staff.^25^

An important finding from this analysis is the differential protection of isoniazid TPT over follow-up. We found that effectiveness was especially high in the first year after exposure (93%) and then dipped to 80% from 1-2 years, and 26% (although not statistically significant), in year 3 and thereafter. To our knowledge, this finding has not been described previously in studies of TPT among close contacts of individuals with tuberculosis. However, it is congruent to results among other vulnerable populations in high-incidence settings where *Mtb* reinfection is thought to occur after sufficient follow-up. For example, among people with HIV from a trial in South Africa, effectiveness of nine months of isoniazid was 48%, 39%, and 12% from <1, 1–2, and after two years of follow-up.^26^ In South Africa, continuous isoniazid was as effective at preventing tuberculosis compared to shorter regimens; however, toleration of the regimen for long durations of time was difficult.^27^ How to manage long-term protection of contacts, including those exposed to drug-resistant disease, is an ongoing discussion.^28^ Despite the waning duration of protection to contacts of multidrug-resistant tuberculosis seen in our study, the observation that protection was high (>80%) for up to 2 years after primary exposure provides important justification to provide TPT in this context.

Our work has limitations. While the dataset is large and from a diverse geographical spread, all studies contributing data were observational. This brings limitations in missing data and potential confounding by indication, given that the choice to give isoniazid was not randomly assigned but decided either at a study level or by clinician decision on an individual basis. For example, we found that a higher proportion of children, who have a higher risk of tuberculosis after exposure than adults, in the isoniazid TPT group.

Although we used propensity score matching to account for differential risk profiling among those receiving and not receiving TPT, residual confounding remains possible. However, if present, this bias is likely to have driven our results towards the null as persons given TPT are likely to have a higher risk profile. Some of the studies contributing data were retrospective. In included studies, differing case definitions were used to define close exposure as well as screening and follow-up for tuberculosis were employed. Some data was not congruently collected across cohorts; therefore, it was not possible to assess the impact of other, potentially important confounders, which may have included cigarette smoking, crowding, socioeconomic status, body mass index, amongst others.

In conclusion, using a combined analysis of nine cohort studies comprising nearly 5,000 close contacts of individuals with MDR tuberculosis, isoniazid substantially and significantly reduced the risk of developing incident tuberculosis, as compared to no treatment. This effect was greater in children than in adults and most apparent in high-burden settings. The duration of protection persisted for two years and then subsequently waned. When developing guidance for MDR-TPT, isoniazid should be considered as a preventive treatment option, alongside fluoroquinolones.

## Data Availability

The data used for this analysis can be made available upon reasonable request to the corresponding author once all relevant substudies from the consortium have been completed.

## Contributors

All authors contributed to data acquisition. LM and JAS conceived the study. LM and JAS were members of the primary writing group and wrote the first draft of the manuscript. LM and JAS edited subsequent versions of the manuscript until dissemination to the broader author group. All authors read and edited the drafted manuscript for important intellectual content and assisted in data interpretation. All authors approved the final version of the manuscript. All authors contributed data to the large, merged individual participant dataset, but some of the data were not accessible to all authors due to data sharing agreements and restrictions by each study group. All authors had final responsibility for the decision to submit for publication.

## Acknowledgments

LM was supported by a National Institutes of Health grant award (1K01AI156022-01). CRH was supported by the Providence/Boston Center for AIDS Research (P30AI042853), the Boston University/Rutgers Tuberculosis Research Unit (U19AI111276) and the Indo-U.S. Vaccine Action Program (VAP) Initiative on Tuberculosis (CRDF Global/ National Institute of Allergy and Infectious Diseases). We acknowledge and thank the participants and investigators in each these studies.

## References

1. Morrison J, Pai M, Hopewell PC. Tuberculosis and latent tuberculosis infection in close contacts of people with pulmonary tuberculosis in low-income and middle-income countries: a systematic review and meta-analysis. The Lancet Infectious Diseases. 2008 Jun 1;8(6):359–68.

2. Martinez L, Shen Y, Mupere E, Kizza A, Hill PC, Whalen CC. Transmission of Mycobacterium tuberculosis in households and the community: a systematic review and meta-analysis. American Journal of Epidemiology. 2017 Jun 15;185(12):1327–39.

3. Coussens A, Zaidi A, Allwood BA, Dewan P, Gray G, Kohli M, Kredo T, Marais B, Marks G, Martinez L, Ruhwald M, Scriba T, Seddon J, Tisile P, Warner D, Wilkinson RJ, Esmail H, Houben RM on behalf of the International Consensus for Early TB (ICE-TB) group. A new disease framework for tuberculosis to guide research towards improved care and prevention - an International Consensus for Early TB (ICE-TB). The Lancet Respiratory Disease. [In-Press]. October 2023.

4. Pai M, Behr MA, Dowdy D, Dheda K, Divangahi M, Boehme CC. Nature Reviews Disease Primers. Tuberculosis. 2016; 2: 16076.

5. Ferebee SH. Controlled chemoprophylaxis trials in tuberculosis. Adv Tuberc Res. 1969; 17: 28–106.

6. Salazar-Austin N, Dowdy DW, Chaisson RE, Golub JE. Seventy years of tuberculosis prevention: efficacy, effectiveness, toxicity, durability, and duration. American Journal of Epidemiology. 2019 Dec 31; 188 (12): 2078–85.

7. Shah R, Khakhkhar T, Modi B, Patel P. Efficacy and Safety of Different Drug Regimens for Tuberculosis Preventive Treatment: A Systematic Review and Meta-Analysis. Cureus. 2023 Apr 26; 15 (4).

8. World Health Organization. WHO consolidated guidelines on tuberculosis: tuberculosis preventive treatment. World Health Organization; 2020 Apr 7.

9. Global tuberculosis report 2023. Geneva: World Health Organization; 2023. Licence: CC BY-NC-SA 3.0 IGO.

10. Knight GM, McQuaid CF, Dodd PJ, Houben RM. Global burden of latent multidrug-resistant tuberculosis: trends and estimates based on mathematical modelling. The Lancet Infectious Diseases. 2019 Aug 1; 19 (8): 903–12.

11. Kherabi Y, Tunesi S, Kay A, Guglielmetti L. Preventive Therapy for Contacts of Drug-Resistant Tuberculosis. Pathogens. 2022 Oct 15; 11 (10): 1189.

12. Seddon JA, Garcia-Prats AJ, Purchase SE, Osman M, Demers AM, Hoddinott G, Crook AM, Owen-Powell E, Thomason MJ, Turkova A, Gibb DM, Fairlie L, Martinson N, Schaaf HS, Hesseling AC. Levofloxacin versus placebo for the prevention of tuberculosis disease in child contacts of multidrug-resistant tuberculosis: study protocol for a phase III cluster randomised controlled trial (TB-CHAMP). Trials. 2018 Dec 20; 19 (1): 693.

13. Fox GJ, Nguyen CB, Nguyen TA, Tran PT, Marais BJ, Graham SM, Nguyen BH, Velen K, Dowdy DW, Mason P, Britton WJ, Behr MA, Benedetti A, Menzies D, Nguyen VN, Marks GB. Levofloxacin versus placebo for the treatment of latent tuberculosis among contacts of patients with multidrug-resistant tuberculosis (the VQUIN MDR trial): a protocol for a randomised controlled trial. BMJ Open. 2020 Jan 2; 10 (1): e033945.

14. Fox GJ, Nguyen CB, Nguyen TA, Nguyen BH, Garden F, Benedetti A, Behr MA, Graham SM, Marais BJ, Menzies D, Marks GB. The effectiveness of lvofloxacin for the treatment of tuberculosis infection among household contacts of patients with multidrug-resistant tuberculosis: the VQUIN MDR trial. 2023 Union Conference on Lung Health. November 2023.

15. Seddon JA, Garcia-Prats AJ, Purchase SE, Osman M, Demers AM, Hoddinott G, Crook AM, Owen-Powell E, Thomason MJ, Turkova A, Gibb DM, Fairlie L, Martinson N, Schaaf HS, Hesseling AC. A phase III cluster randomised controlled trial to assess efficacy of preventive treatment in child contacts of multidrug-resistant tuberculosis. 2023 Union Conference on Lung Health. November 2023.

16. Huang CC, Becerra MC, Calderon R, Contreras C, Galea J, Grandjean L, Lecca L, Yataco R, Zhang Z, Murray M. Isoniazid preventive therapy in contacts of multidrug-resistant tuberculosis. American Journal of Respiratory and Critical Care Medicine. 2020 Oct 15;202(8):1159–68.

17. Martinez L, Cords O, Horsburgh CR, et al. The risk of tuberculosis in children after close exposure: a systematic review and individual-participant meta-analysis. The Lancet. 2020 Mar 21;395(10228):973–84.

18. Martinez L, Cords O, Liu Q, et al. Infant BCG vaccination and risk of pulmonary and extrapulmonary tuberculosis throughout the life course: a systematic review and individual participant data meta-analysis. The Lancet Global Health. 2022 Sep 1; 10 (9): e1307–16.

19. Martinez L, Seddon JA, Horsburgh CR, Lange C, Mandalakas AM, TB Contact Studies Consortium. Effectiveness of preventive treatment among persons with differing age and Mycobacterium tuberculosis infection status: A systematic review and individual-participant meta-analysis of contact tracing studies. The Lancet Respiratory Health. [Online Only]. March, 2024

20. Teixeira L, Perkins MD, Johnson JL, Keller R, Palaci M, do Valle Dettoni V, Canedo Rocha LM, Debanne S, Talbot E, Dietze R. Infection and disease among household contacts of patients with multidrug-resistant tuberculosis. The International Journal of Tuberculosis and Lung Disease. 2001 Apr 1;5(4):321–8.

21. Sneag DB, Schaaf HS, Cotton MF, Zar HJ. Failure of chemoprophylaxis with standard antituberculosis agents in child contacts of multidrug-resistant tuberculosis cases. Pediatric Infectious Diseases Journal. 2007;26:1142–1146.

22. Seifert M, Catanzaro D, Catanzaro A, Rodwell TC. Genetic mutations associated with isoniazid resistance in *Mycobacterium tuberculosis* : a systematic review. PloS One. 2015 Mar 23;10(3):e0119628.

23. Lempens P, Meehan CJ, Vandelannoote K, Fissette K, de Rijk P, Van Deun A, Rigouts L, de Jong BC. Isoniazid resistance levels of Mycobacterium tuberculosis can largely be predicted by high-confidence resistance-conferring mutations. Scientific Reports. 2018 Feb 19;8(1):3246.

24. Khan SR, Manialawy Y, Siraki AG. Isoniazid and host immune system interactions: A proposal for a novel comprehensive mode of action. Br J Pharmacol. 2019 Dec;176(24):4599–4608.

25. Schluger NW. Prevention of Multidrug-Resistant Tuberculosis in Close Contacts. Back to the Future?.American Journal of Respiratory and Critical Care Medicine. 2020 Oct 15;202(8):1077–8.

26. Rangaka MX, Wilkinson RJ, Boulle A, Glynn JR, Fielding K, Van Cutsem G, Wilkinson KA, Goliath R, Mathee S, Goemaere E, Maartens G. Isoniazid plus antiretroviral therapy to prevent tuberculosis: a randomised double-blind, placebo-controlled trial. The Lancet. 2014 Aug 23; 384 (9944): 682–90.

27. Martinson NA, Barnes GL, Moulton LH, Msandiwa R, Hausler H, Ram M, McIntyre JA, Gray GE, Chaisson RE. New regimens to prevent tuberculosis in adults with HIV infection. New England Journal of Medicine. 2011; 365: 11–20.

28. Churchyard G, Cárdenas V, Chihota V, Mngadi K, Sebe M, Brumskine W, Martinson N, Yimer G, Wang SH, Garcia-Basteiro AL, Nguenha D. Annual tuberculosis preventive therapy for persons with HIV infection: a randomized trial. Annals of Internal Medicine. 2021 Oct;174(10):1367–76.

