## Supplement for "The effectiveness of isoniazid preventive treatment against tuberculosis among contacts of multidrug-resistant tuberculosis: A systematic review and individual-participant meta-analysis"

**Tables and Figures.**

Additional Methodological Information.

Table S1. Demographic characteristics of included cohort studies

Table S2. Study results with information on multi-drug resistant status of index cases.

Table S3. Effectiveness of isoniazid preventive treatment against incident tuberculosis among contacts exposed to MDR tuberculosis cases, by time period of follow-up.

Table S4. Incidence rate ratios among subgroups of children 0 exposed to multidrug-resistant tuberculosis, stratified by the age of the child.

Table S5. Incidence rate ratios among subgroups of participants exposed to multidrug-resistant tuberculosis, stratified by the background tuberculosis incidence of the country.

Table S6. Assessment of Quality of the Included Studies.

Table S7. Demographic characteristics of participants administered and not administered preventive treatment.

Figure S1. Map of Location of Included Studies.

Figure S2. Two- and four-year follow-up for tuberculosis, stratified by isoniazid preventive treatment given

**Additional Methodological Information.**

Further enrollment and follow-up from original consortium data

Following the publication of the manuscript Anger et al, 2012, the New York City Department of Health continued to expand their dataset as part of routine programmatic activities. Contact tracing and follow-up data from 2007 to 2019 were added to the New York City cohort for the purposes of this analysis. Although contact tracing was done by the same group (New York City Department of Health), exact methods throughout this time period are not exactly the same. For example, IGRAs were used in later years of the study time period.

Event Ascertainment

Events were ascertained using several strategies selected by each cohort's investigator group. For tuberculosis diagnosis events, cohorts either diagnosed cases prospectively or used data linkage to national or sub-national tuberculosis registries. Most prospective studies used some type of microbiological test, either as a baseline evaluation or a triage test. Diagnosis of tuberculosis in all prospective studies included either a positive microbiological test or a physical examination and subsequent clinical diagnosis. Some prospective studies used diagnostic tests as part of their study procedures and also used national or sub-national tuberculosis registries.

Systematic Search.

Case-control studies and outbreak reports were excluded, as were reviews, editorials, letters, or studies for which individual outcomes were not reported. We did not restrict articles by language and reviewed manuscripts written in English, Chinese, French, German, Japanese, Korean, Persian, Portuguese, Russian, and Turkish. To facilitate the review process, a list of 'exclusionary words' was developed, based on words in titles highly suggestive of irrelevant content (see below for complete explanation and list). Manuscripts were excluded if their titles contained any of these exclusionary words. Two reviewers independently reviewed articles in two stages: evaluation of titles and abstracts followed by full-text review. After the review of titles and abstracts, the two reviewers discussed discrepancies and re-evaluated articles until consensus was reached. During this stage, if an abstract was in a language other than English the manuscript was advanced to the full-text review stage. Relevant articles were subject to a full-text review by both reviewers and any discrepancies were again resolved by reviewer discussion and consensus. If eligibility could not be assessed from the full-text manuscript because of missing information, we contacted authors for clarification. We evaluated eligible articles for duplication of data on the same individuals and excluded manuscripts if necessary.

Preventive Treatment

Preventive treatment was assigned to participants according to each study's protocol or local guidelines and practices.

Study Quality.

Each study was judged based on a 9-point scale using three broad criteria: selection of participants (4 points), comparability of studies (2 points), and ascertainment of outcome of interest (3 points). High study quality was defined as >66.6%, moderate quality as 33.3-66.6%, and low quality as <33.3%. Discrepancies between the two reviewers were resolved by re-evaluating the study for consensus.

Study-level characteristics.

Country-level tuberculosis incidence data was collected from World Health Organization databases for each study. This variable was used as a continuous variable. Studies were categorized into "high-burden" country as classified by the World Health Organization. Studies were also grouped into World Health Organization global regions and World Bank country-level economies (high- income, upper-middle-income, lower-middle-income, and low-income) as of October 2018.

**Search Strategy.**

**Pubmed/MEDLINE – 6252**
Search conducted on April 7, 2018
(tuberculosis OR (mycobacterium tuberculosis[MeSH Terms]) OR tuberculosis[MeSH Terms])
AND
(contact tracing[MeSH Terms] OR infectious disease contact tracing[MeSH Terms] OR household*[Title/Abstract] OR contact*[Title/Abstract])

**Embase – 8525**
Search conducted on April 7, 2018
(tuberculosis OR 'mycobacterium tuberculosis')
AND
('contact examination'/de OR contact*)

**Biosis – 3971**
Search conducted on April 7, 2018
((TS=tuberculosis) OR (TS='mycobacterium tuberculosis') OR (TI=TB))
AND
((TS='contact examination') OR (TS=contact*) OR (TS='contact tracing') OR (TS=outbreak*))

**Web of Science – 6537**
Search conducted on April 7, 2018
((TS=tuberculosis) OR (TS='mycobacterium tuberculosis') OR (TI=TB))
AND
((TS='contact examination') OR (TS=contact*) OR (TS='contact tracing') OR (TS=outbreak*))

<u>Total from Search:</u> 25285.

<u>Total from Search after Exclusion by Duplicates:</u> 14927

<u>Total from Search after Exclusion by Timeframe (pre-1998):</u> 9753

**Exclusionary Keyword Algorithm for Study Titles.**

i. Description of the Algorithm.

We wrote a script parsing titles into individual words. We selected words highly suggestive that an article was unrelated to our study objectives. Articles were then eliminated based on whether their titles contained these words.

In order to validate this process, we implemented the algorithm on the first 100 titles and manually screened them for eligibility in the study. Our exclusionary algorithm eliminated all articles that were screened out by manual screening with 100% specificity. This suggested that our selected words were appropriate. Out of the 9,243 articles disqualified at the title stage, 1,829 (19.7%) were eliminated based on whether they contained one of the exclusionary words. The code for exclusion words and article elimination, as well as the entire list of exclusionary words can be found below.

ii. Python script:

```
'''
Script parsing tokens for excluding titles and matching the tokens to the original word counterpart to each token.

Each title is normalized to lower-case letters and numbers with no punctuation. Excluding tokens use this format.
A list of exclusion tokens is used to eliminate all matching titles.
'''
import pandas as pd
import string
from difflib import SequenceMatcher

import sys
reload(sys)
sys.setdefaultencoding('utf8')

def remove_punctuations(text):
    """Removes punctuation from the string text.

    :param text: string
    :return: string, with punctuation removed.
    """
    if isinstance(text, float): return text
    for punctuation in string.punctuation:
        text = text.replace(punctuation, '')
    return text

def similar(a, b):
    """
    Returns ratio of similarity between a and b in the range [0, 1].
    :param a: string
    :param b: string
    :return: float, between 0 and 1 inclusive
```

```
    """
    return SequenceMatcher(None, a, b).ratio()


def find_ex_word(ex_words_set, r):
    """
    Matches title words to exclusionary words.

    :param ex_words_set: set<string> of exclusionary words.
    :param r: pandas row
    :return: [(fraction, exclusion_word, title_word)] or 'No match' for no matches. The list contains all possible
matches.
    """
    candidates = []
    if isinstance(r['Title_punctuations'], float):
        return 'No match'
    for word in r['Title_punctuations'].split(' '):
        if word in ex_words_set:
            for original in r['Title'].split(' '):
                candidate = (similar(word, original.lower()), word, original)
                candidates.append(candidate)
    candidates.sort(key=lambda x: x[0], reverse=True)
    candidates = [c for c in candidates if c[0] > 0.0]
    if not candidates:
        return 'No match'
    else:
        return candidates

def run():
    '''Main driver function '''
    all_articles_read = pd.read_csv('endnote_oliviasearch_0411.csv')  # read File containing titles
    all_articles_read = all_articles_read.dropna(subset=['Title'])  # remove blanks
    all_articles_read['Title_punctuations'] = all_articles_read['Title'].apply(remove_punctuations)
    all_articles_read['Title_punctuations'] = all_articles_read['Title_punctuations'].str.lower()  #
make lowercase
    title_word_count =
(all_articles_read['Title_punctuations'].str.split(expand=True).stack().value_counts(
        ascending=True))  # list of title with corresponding number of occurrences

    # Sheet of words and counts across titles.
    (pd.DataFrame(title_word_count).to_excel('title_word_count.xlsx', index=True))  # export
title_word_count

    # Read in file of exclusion words. Filter out titles that contain them.
    exclusion_words = pd.read_excel('exclusion_words.xlsx')  # read File containing words to exclude

    exclusion_word_delim = [r'\b' + item + r'\b' for item in exclusion_words.word]
```

```
    exclusion_words_str = '|'.join(exclusion_word_delim)
    filtered = all_articles_read[all_articles_read['Title_punctuations'].str.contains(
        exclusion_words_str) == False]  # remove article titles if they contain exclusion words
    (pd.DataFrame(filtered).to_excel('907_exclude_test.xlsx', index=True))  # export remaining titles

    ex_words_set = set()

    for word in exclusion_words['word']:
        ex_words_set.add(word)

    # List of tuples of match ratio (0-1), token, original_word
    matches = all_articles_read.apply(lambda x: find_ex_word(ex_words_set, x), axis=1)

    # build a dictionary from token to the best possible match
    ex_dict = dict()
    for match in matches:
        if match != 'No match':
            for n in match:
                key = n[1]
                if key not in ex_dict:
                    ex_dict[key] = n
                else:
                    ex_dict[key] = max(n, ex_dict[key])

    # mapping from token to original word
    matched_dict = dict()
    for value in ex_dict.values():
        if value[0] > 0:
            matched_dict[value[1]] = value[2]

    # see if any of the exclusion words were never matched to an original title word
    words_not_found = []
    for w in ex_words_set:
        if w not in matched_dict:
            words_not_found.append(w)

    original_words = [x[2] for x in ex_dict.values()] + words_not_found # write all words out
    serialized = ', '.join(original_words)
    f = open('/Users/ocords/Desktop/original_words_test.txt', 'w')  # creates text files words as
appear in original titles
    f.write(serialized)
    f.close()


if __name__ == '__main__':
    run()
```

iii. List of words:
anti-coronavirus, A(2)CoMnO(6), Sulfhydrylase, influenza, polypeptides, lysosome, Thermococcus, YMn6-xTixSn6, amphotericin, reductoisomerase, porcine, Enteric, cryptococcosis, milk, cytokines, langurs, perianal, Birthplace, benzoquinone:, dichloroacetate, penile, polyunsaturated, Protein-RNA, peanut, squirrels:, Halogen, dairy, 3-(pyrazin-2-ylcarbonyl)dithiocarbazic, choliangiopancreatography, leukocytes, rhesus, ppe38, dermatophytosis, heme, Biofilms, extremophiles, UDP-galactopyranose, D-3-phosphoglycerate, peafowl, Salmonella, monkeys, ESAT-6-dependent, O-Acetylserine, mongooses, Primate, spoligofamily, (+1188A/C), Nicotinamide, vitrectomy, Isoniazid/Rifampicin/Poly, Shiga, Trypanosoma, A/H3N2, brucellosis, veterinarians, wild-boar, phosphorylated, Amoeba-Resistant, Cyclodestructive, pJHCMW1, miliary, subspecies, prostatitis, enterocolitica, hydrolysis, hematology-oncology, prisons, histone-like, macrophages, vampire, Pseudoaneurysm, lovebird, celiac, Guanine-Cytosine-Rich, phosphatase, Hsp70, factor-microRNA, osteoporosis, bovis, Bird, S12-S7, actinobacterial, CRF08_BC]., Corynebacteriurn, raccoons, brucei, ribose, kangaroo, herd, PD-1/PD-L2, beta-cyclocitral, alphaA-crystallin,, water-soluble, Ag85B, PCR-restriction, helicase, CXCL10/IP-10, inter-species, beta-semialdehyde, Asp299Gly, host-parasitoid, corynebacterium, Paracoccidioides, cinnamon,, Orang-Utan, metal-induced, raccoon, military, lymphadenitis, calf-to-calf, Lanthanide(III)-Phthalocyanine, sacroiliitis, Pseudomonas, extremophile:, mct1Delta, MD-2, Anions, glutaraldehyde, earth-nickel-indides, Coxsackievirus, transplant, arthropod, obliterans, catalase-peroxidase, Dy5Ni2In4, petroleum, Frog, oryx, low-molecular-mass, Automata, microbiota, electroelution, phytopathogen, (P631H), IL-17RA, psoriasis, Shiga-toxin-producing, staphylococcus, Microaggregates, 49-year-old, SAT6-CFP10, E-coli, chimpanzee, Metalloprotease-1, Adenosine, biotin, rabbit, radiculomyelitis, penis, Rv0753c, flora, animals, GC1237, K182G, PstS-1(285-374):CFP10, Death-Ligand, cat, mammalian, Arg753Gln, oxide, ligase, MDP-1, C1858T, proteasomal, Helicobacter, aminoglycoside, neurosarcoidosis, nursing, gingiva., Rv1737c, ferrets, pelvic, camelids, keratitis, CYP121-fluconazole, alpacas, gingivalis, cows, Oligosaccharides, confocal, hydrogen, species, Lamb, ribokinase, Lumbricidae), jails, paleopathological, pharyngitis-a, Cryoannealing-induced, rhinoceros, Micelle-based, animal, elephant, oropharyngeal, goat, ligand-independent, fever, aureus, Bacteriophages:, canker, fungus, possum, pigs, M2e.HSP70c, MVA85A,, prostate:, Leishmania, Bloodstream, calves, immune-endocrine, macrolide,, crystallin, disposable-sheath, jail, methadone, ID83/GLA-SE, Channel-Forming, crystallographic, Association-of-Primate-Veterinarians,, CD127-cells, antelope, phagocytosis, cryptosporidiosis, albicans, RE4Ni11In20, ulcerans, epilepsy, brucellosis--a, avium, PD-Ligand, Game-Theoretic, nitrogen, gonadal, Carnivores, 1-deoxy-D-xylulose-5-phosphate, badger, FMO2, leprae, antigen/N-trimethylaminoethylmethacrylate, ligand, botanical, protein-3, transferase, Cryptosporidium, m(1)A58, flavins, Rv1735c,, conspecific, Foxp3, coffee., cholera, food, diphosphate, osteolytic, Tb-2(SO4)(3), hypoxic, chemokines, phagocytes, exon, (Giraffa, metabolite, nematodes, DT104, ClpP1P2,, meat, aeruginosa, thymus., hemagglutinin, neoformans, esat-6, phytopathogenic, coronavirus, 33-year-old, amphiphilic, pyrimidine, CFP21-MPT64, bed-nets, sulfoglycolipids, D543N, chlamydial, Animal-Derived, myelin, elephants, Enterobacteriaceae, fish, sugars, bovine, 11p14-15, oncological, Quantum, lysis, protein-II, C(-159)T, semen, heme-degrading, metagenome, MPT51, phosphoryl, CD8+T, wildlife-pathogen, anorexia, (sIL-7R), cyber-gaming, c.1770-1900, arthritis, zoological, alkynes., lipopolysaccharide-induced, O3157, oligonucleotide, pre-ulcer, mammal, substrate-binding, host-microbial, agarose, monkey, phage-based, equine, SLC11A1, H1N1, braziliensis, Pseudo-septic, ostrich, Opiate-Driven, G354R, swine, Cyanide, osteomyelitis, phagosomes, ionic, raptors, borreliosis, sympatric, helix-turn-helix, buffaloes, leishmaniosis, 6-kilodalton, small-bowel, malonyl-CoA:AcpM, exostosis:, TLR4, antirheumatic, mammary, Earthworms, b-cell, sheep, ESAT-6/CFP-10, RMn6X6-x, pyrophosphatase., leptospirosis, sapiens,

chimaera, Variable-Number-Tandem-Repeats, fragment-length, dyskinesia, leprae-specific, 4-Sulfamoylphenyl-omega-aminoalkyl, amines, pylori-Associated, Variant-Repeat, Carotenoids, human-livestock-wildlife, cytokine--IL-12,, resuscitation-promoting, Endoluminal, rickshaw, inmates, herds, Caulobacter, pheasants, non-contact-lens, vulvar, hemangiopericytoma:, mannose-binding, H-2K(k), IS6110, microglial, vulval, IL1B, flavin-binding, apiospermum, tubules, A0248:, mink, macaw, Legionella, fowl, kDa/MPT-64,, phosphoantigen-mediated, foodborne, phosphoribosyltransferase, lymphoblastic, rat, menstruus), dendritic, CD4+CD25, IS6110-fAFLP, glycopeptidolipid, MyD88, 30/31-kDa, smallpox, aeruginosan, amphibious, Crystallography, Tattoo-Related, clonality, DNA-probes, Xanthium, zoo, endangered, heat-shock, bullfrogs, serine, HLA-A*0201-Restricted, Chromosomes, botulinum, EGGS., protein-10, cytokines/chemokines, free-ranging, host-parasite, Hsp16.5, carcinoma, Brucella, vitro-selected, Elk-Fetus, ranavirus, Rv0081,, hemothoraces, Adenine, minisatellites, hydrocephalus, phospholipase, mannose, kDa/CFP-10,, socioepidemiologic, H-2-rich, fungal, bioarchaeology, cholerae, coli, human-wildlife, macaque, Rv1498A., pseudodiphtheriticum, aviary, ulcer, 30-kDa, hemangioendothelioma-case, (RE)(12)Co5Bi, G2109A), bronchoscopy-a, camelopardalis), kinase, GlfT2,, otorhinolaryngology, ESAT-6/MPT-64, Wood, Demodecidosis, salmonellosis, N-acetyl-gamma-glutamyl-phosphate, glycolipid, slit-lamp, Methylisothiazolinone, Nanocluster, palaeopathological, (10.1016/S1473-3099(17)30447-4)), histone, asbestos, Orang, endocarditis:, Cytokine-based, cyclopropane, cervids, cj0183, chrysomya, primates, CD8, Campylobacter, sarcoidoisis, CD14-159C/T, spondyloarthritides, KIR3DL1/S1:, faeciuml,d-transpeptidase, abortus, dUTPases, tumors, heterocyclic, O157:H7/H-strains, chitotriosidase, lymphokine, IL-12Rbeta2, PENGUINS, bison, cyanobacterial, Hydrogen-bonding, aegypti, Synechococcus, pseudokinase, uveitis, 10.1093/cid/ciw694), Thymidylyltransferase, alanine, supramolecular, L-isoleucine, anthropozoonotic, chondrosarcoma, 2,3-Naphthalocyaninato, ionization-time-of-flight, CD1d-dependent, cattle, Salmonellae, thrombocytopenic, thrombocytopenia, IL12RB1, Neurobrucellosis:, House-Roosting, CHIMPANZEES, nucleoside, pets, zebrafish, ebola, CD1-mediated, chaperonin, (S1473309917304474), CD8-positive, gonorrhoeae, lymphocyte, (T874A, Clavibacter, Chihuahua, zoonoses, bushbuck, cervical, exomes, STAT3, Cow, hantavirus, cruzi, gyrase-fluoroquinolone, peptidoglycan, androgens, filariasis., CCR4, single-amino-acid, Ms6564,, lipases, Arg677Trp,, Tattoo-associated, Dysphagia, cancer, CRISPRs, Synthases,, Cryptogenic, peptide-binding, Clostfdium, thermophilus, crystalline, grazing, amino, myeloid-derived, Rv3802c, beef, Enterococci, (-362g/c), intracellular, benzaldehydes:, tracheobronchopathia, macaques, chromatography-tandem, interleukins, hydrolase, SrCl2-Promoted, pseudogene, Rv1057, isomerase, CD41, Lipid-Polymer, chain-binomial, CD40, Hydrogels, 1,2,4-triazoles,, ophthalmic, actinomycetemcomitans, Otolaryngological, miR-26a,, species-history, Apoptosis-associated, (S0140673615001518), tortoise,, cytotoxic, MazF-mt6, haematobium, Toxoplasma, Herpesvirus, CYP2E1, MS0006, leprosy, wild, cell-entry, interleukin-4, galactofuranosyltransferase, interleukin-1, cell-wall, CD1-presented, methyltransferases, glycaemic, Strongyloidiasis:, PstS1(285-374):CPF10:, ducks, polymerase, Proteolytic, eczema, sarcoma, beta-Lactamases:, Upregulated, 5-Phosphate, Nonsyphilitic, (CD11b/CD18), hyodysenteriae, ribosome, larva, Ccr2, helicases, Dengue/Zika, palsies, rrs491, cytolysis, Chromolaena, proleukin, IS3-based, chickenpox, Neutron, alcohol-resistant, zoonosis, 3-Deoxy-D-manno-octulosonate, difficile-associated, gastroenterology, nicotinohydrazide, typhoid, Anesthesiology, streptococcus, epizooties, ML2331, transposon, hydroxylase, poultry, 3-dioxygenase, C-24-methyltransferase, cytosolic, Th1/Th2, (IL)-12p70, chickens, vertebra:, chemokine, giraffe, post-implantkeratoprosthesis, 8-Phosphate, brasiliensis, streptomycin, lambs, mechanism-based, biochip, livestock, Glycine, ORS571, Lentivirus-control, Carboxylic, scabies, super-oxidized, mutase, sarcoidosis, Chagas, Mongoose, dpp3, macroscopic, Pyrazinamidase, cervix, boars, catalytic, RD1-epitopes, ML1419c, swine-origin, papulonecrotic, lattice, matricellular, prison, lymphocytes, nitrocellulose-bound, murine, IL-17-producing, parenchymal, apnea., scrofuloderma, nanofilters, prosthesis-free, Ospedalieri., wavelength, '-[(E)-2,6-Dichlorobenzylidene]pyrazine-2-carbohydrazide, C8G, Dy(III)-Phthalocyanine, furanoside, lymphoma, bioterrorism-related, Glucose-1-Phosphate, Sulfonyl-hydrazones, glycolipid-I, qualitative, spelunking:, brachiocephalic, Kuala, nosocomial, Estriol, Amoebae, antiprotozoal, leucoryx,, partridges, CeFeSi-type, ulcers, EMRSA-15, 38-kDa, CD56+CD3+, CYP121:, (H5N1), Protein-Ligand, CXCL10, Transcriptome, Gonococcal, pestis, gastrostomy, ungulate, pet, veterinary, fox, anti-leishmanial, goats, e59414,, syphilis, H7N9, wild-caught, zoonotic, IS6110-RFLP, metal/metal, core, Rv2721c), Tetrakis(4-chlorophenyl)borate, abattoirs, squirrel, CFP10, CCL18, dental, p38, autophagosomes, nanoliter, leukocyte-leukocyte, tetramer, wildlife, Autophosphorylation, Rv2628, tumor, Lynx, tannic, alpha-Substituted-2-Phenylcyclopropane, ribose-5-phosphate, Posttraumatic, badgers, pH-dependent, CYP51., aortic, protease, F15/LAM4/KZN, haplotype, D2EHPA, (pyrazinecarbonyl)hydrazones, mitochondrial, sow., ex-vivo, non-ruminant, dehydrogenase, MCP-2, neutrophil-mediated, larvae., reticulum-related, crystallization, herbivores, Larval, Oligomycin, polymerization, cats, nitric, PPE39, 5q31.1, ML0405, CD1-lipid, CD4+CD45RO+T-Cells, tyrosine, metaproteomics., helminths, Convex-probe, chikungunya, mammals, keratinocyte, chromosomal, vivax, PCC7942., phenylalanine, Zika, PTPN22, CYP125:, Enterovirus, malate, Peppermint, apes, Octahydrocyclopenta[c]pyrrol-2-yl, aneurysm, 38kDa-antigen, polymorphisms, 'Zebra', rabbits, Cpn60.2, kinetics, Cytokine-Induced, deer, Enterococcus, Rv1733c,, pig, demethylase, lichen, Hypercalcemia, ticks, NOS2A, CFP10ESAT6, Rv0183, 14alpha-sterol, Lgn1, tandem-repeat, keratoplasty, Coxiella, N-(4-Bromophenyl)pyrazine-2-carboxamide, peptides, UVB-irradiated, snakes, photoluminescence, nanopore, thromboendarterectomy, actinorhodin, Glycosaminoglycans, IS1106, arthritis-associated, carboxylase, Vibrio, diesters, N-(2-Chloroethyl)pyrazine-2-carboxamide, peptide-25, inguinal, macrophage, non-methylated, cellulose-targeting, PCR-single-strand, camelus)., Flavohemoglobin, RegX3, Neuropilin-1, gonorrhea, Bartonella, citrate, electroretinographic, tnf_il2, bovus, antitnf_, v_11, guinea pig, 5_untranslated

**References for All Individual Studies in Analysis.**


Altet, N., Dominguez, J., Souza-Galvão, M.L.D., Jiménez-Fuentes, M.Á., Milà, C., Solsona, J., Soriano-Arandés, A., Latorre, I., Lara, E., Cantos, A. and Ferrer, M.D., 2015. Predicting the development of tuberculosis with the tuberculin skin test and QuantiFERON testing. *Annals of the American Thoracic Society*, 12(5), pp.680-688.

Anger, H.A., Proops, D., Harris, T.G., Li, J., Kreiswirth, B.N., Shashkina, E. and Ahuja, S.D., 2012. Active case finding and prevention of tuberculosis among a cohort of contacts exposed to infectious tuberculosis cases in New York City. *Clinical Infectious Diseases*, 54(9), pp.1287-1295.

Chakhaia, T., Magee, M.J., Kempker, R.R., Gegia, M., Goginashvili, L., Nanava, U. Blumberg, H.M., 2014. High utility of contact investigation for latent and active tuberculosis case detection among the contacts: a retrospective cohort study in Tbilisi, Georgia, 2010–2011. *PloS One*, 9(11), p.e111773.

Chan, P.C., Shinn-Forng Peng, S., Chiou, M.Y., Ling, D.L., Chang, L.Y., Wang, K.F., Fang, C.T., Huang, L.M., 2014. Risk for tuberculosis in child contacts. Development and validation of a predictive score. *American Journal of Respiratory and Critical Care Medicine*, 189(2), pp.203-213.

Grandjean, L., Gilman, R.H., Martin, L., Soto, E., Castro, B., Lopez, S., Coronel, J., Castillo, E., Alarcon, V., Lopez, V. and San Miguel, A., 2015. Transmission of multidrug-resistant and drug susceptible tuberculosis within households: a prospective cohort study. *PLoS Medicine*, 12(6), p.e1001843.

Grandjean, L., Crossa, A., Gilman, R.H., Herrera, C., Bonilla, C., Jave, O., Cabrera, J.L., Martin, L., Escombe, A.R. and Moore, D.A.J., 2011. Tuberculosis in household contacts of multidrug resistant tuberculosis patients. *The International Journal of Tuberculosis and Lung Disease*, 15(9), pp.1164-1169.

Hannoun, D. and Boulahbal, F., 2016. Incidence of tuberculosis among children living in contact with smear-positive tuberculosis: Advantages and limits of the Quantiferon TB gold in tube test. *International Journal of Mycobacteriology*, 5, p.S3.

Lu P, Ding X, Liu Q, Lu W, Martinez L, Sun J, Lu F, Zhong C, Jiang H, Miao C, Zhu L. Mediating effect of repeated tuberculosis exposure on the risk of transmission to household contacts of multidrug-resistant tuberculosis patients. *The American Journal of Tropical Medicine and Hygiene*. 2018 Feb; 98 (2): 364.

Mazahir, R., Beig, F.K., Ahmed, Z. and Alam, S., 2017. Burden of tuberculosis among household children of adult multi drug resistant patients and their response to first line anti tubercular drugs. *Egyptian Pediatric Association Gazette*, 65(4), pp.122-126.

Table S1. Demographic characteristics of included cohort studies

| Study or participant characteristics | Studies (n=9) | Proportion |
|---|---|---|
| Prospective study design | 5 | 56 |
| World Health Organization high-burden country* | 2 | 22 |
| Background burden, >100 incident cases per 100 thousand people | 3 | 33 |
| Country-level tuberculosis incidence (per 100 000 people)† | | |
| Below 50 | 2 | 22 |
| 50–100 | 4 | 44 |
| >100–200 | 2 | 22 |
| Above 200 | 1 | 11 |
| World Health Organization region | | |
| African | 1 | 11 |
| Americas | 3 | 33 |
| Eastern Mediterranean | 0 | 0 |
| European | 2 | 22 |
| South-East Asia | 1 | 11 |
| Western Pacific | 2 | 22 |
| HIV status of participant reported | 5 | 56 |
| Cohort size | | |
| Less than 1000 | 7 | 78 |
| 1000 or greater | 2 | 22 |
| QuantiFERON or tuberculin skin testing | 7 | 78 |

Table S2. Study results with information on multi-drug resistant status of index cases.

| Characteristic | $N_{treated}/N_{total}$ | Adjusted Hazard Ratio (95% CI) |
|---|---|---|
| All participants (Model 1) | 722/4,945 | 0.41 (0.23-0.72) |
| All participants (Model 2) | 722/4,945 | 0.32 (0.18-0.58) |
| All participants (Model 3) | 722/4,945 | 0.30 (0.16-0.56) |
| Age group, years | | |
| Child, <18 | 583/1,467 | 0.39 (0.18-0.87) |
| Adult, 18 and over | 139/3,425 | 0.44 (0.14-1.41) |
| HIV status | | |
| Positive | 5/329 | Undefined |
| Negative | 610/3,271 | 0.29 (0.16-0.54) |
| Prior tuberculosis | | |
| Yes | 11/313 | 0.55 (0.13-2.36) |
| No | 653/4,227 | 0.34 (0.16-0.69) |
| TST or IGRA status | | |
| Positive | 134/954 | Undefined |
| Negative | 33/664 | Undefined |

Model 1 includes all contacts and is adjusted for age and the study design and has a random study-level effect.
Model 2 includes all contacts and is adjusted for age, sex, the study design and has a random study-level effect.
Model 3 includes contacts and is adjusted for age, sex, prior tuberculosis, the study design and has a random study-level effect.

Table S3. Effectiveness of isoniazid preventive treatment against incident tuberculosis among contacts exposed to MDR tuberculosis, by time period of follow-up.

| | $N_{events}$ | **Person-years** | **aHR** | **95% CI** |
|---|---|---|---|---|
| Time period | | | | |
| <365 days | 70 | 4,712 | 0.07 | 0.02-0.52 |
| 365 to 729 days | 35 | 3,561 | 0.20 | 0.05-0.89 |
| 730 days and after | 48 | 2,749 | 0.74 | 0.34-1.59 |

aHR: adjusted hazard ratio

Table S4. Incidence rate ratios among subgroups of children exposed to multidrug-resistant tuberculosis, stratified by the age of the child.

| | **Incidence rate ratios** | **95% confidence interval** |
|---|---|---|
| Age, years | | |
| 0 to 4 | Undefined** | |
| 5 to 9 | 1.93 | 0.39-9.59 |
| 10 to 19 | 0.44 | 0.18-1.05 |

** 0 events occurred among 176 children in this category that took isoniazid TPT. Among 247 children in this category that did not take TPT, five (2%) developed incident tuberculosis. Due to no events in the isoniazid TPT group, a measure of association was not able to be calculated.

Table S5. Incidence rate ratios among subgroups of participants exposed to multidrug-resistant tuberculosis, stratified by the background tuberculosis incidence of the country.

| | **Given INH TPT** | | **Not given TPT** | | **Risk difference (95% CI)** |
|---|---|---|---|---|---|
| | **$N_{participants}$** | **$N_{events}$** | **$N_{participants}$** | **$N_{events}$** | |
| Background incidence (per 100K persons) | | | | | |
| <50 | 135 | 0 | 854 | 10 | 1.1 (0.4, 1.9), $P_{exact}$=0.61 |
| 50 to 100 | 36 | 0 | 729 | 1 | 0.1 (-0.1, 0.4), $P_{exact}$ =1.00 |
| >100 | 591 | 13 | 2,595 | 127 | 2.7 (1.2-4.1), P=0.0047 |

Table S6. Assessment of Quality of the Included Studies.

We assessed the quality of each study based on a modified version of Newcastle-Ottawa scale (Wells, 2012)†. We assessed each study's selection process, comparability, and outcome for a maximum total of 9 points. Studies were ranked high if they had a score of greater than 66.6%, moderate if they had a score greater than 33.3 and less than or equal to 66.6%, and low if they had a score of less than or equal to 33.3%.

| **Study†** | **Selection** | | | **Comparability** | **Outcome** | | | **Rating** | | **Quality** |
|---|---|---|---|---|---|---|---|---|---|---|
| | **Represent-ativeness of sample[a]** | **Ascertainment of exposure status - How was index case diagnosed?[b]** | **Demonstration that TB in contacts was not present at baseline. [c]** | **Comparability of cohorts (2 points)[d]** | **Assessment of tuberculosis (2 points)[e]** | **Was follow up long enough for outcomes to occur?[f]** | **Adequacy of follow up of exposed contacts [g]** | **Rating** | **PERCENT SCORE (X/9)** | |
| Grandjean, 2010 | A* | A* | C | B*/C | A*/C | A* | E | 5 | 0.56 | moderate |
| Grandjean, 2015 | C | A* | A* | A*/B* | A*/B* | A* | E | 7 | 0.78 | high |
| Chan, 2014 | A* | A* | A* | B*/C | A*/B* | A* | E | 7 | 0.78 | high |
| Chakhaia, 2014 | C | A* | A* | C/D | A*/B* | A* | E | 5 | 0.56 | moderate |
| Altet, 2015 | A* | A* | A* | A*/B* | A*/B* | A* | B* | 9 | 1.00 | high |
| Mazahir, 2017 | A* | A* | A* | A*/D | A*/B* | A* | A* | 8 | 0.89 | high |
| Lu, 2015 | B* | A* | B* | B* | D | A* | C* | 6 | 0.67 | moderate |

| | | | | | | | | | | |
|---|---|---|---|---|---|---|---|---|---|---|
| Anger, 2012 | B* | A* | A* | C/B* | B* | A* | C* | 7 | 0.78 | high |
| Gounder, 2015 | A* | A* | A* | C/B* | B* | A* | C* | 7 | 0.78 | high |

†Hannoun, 2016 was a conference abstract and therefore was not included in the assessment.

[a] A*- Representative of the average TB case/TB contact in the community; B*- Somewhat representative of the average TB case/TB contact in the community; C - Selected group of TB cases/TB contacts, chance of bias; D - No description of the derivation of the cohort;
[b] A*- Microbiological (smear, culture, Xpert) testing of TB cases was done for all tuberculosis index cases; B - Chest radiographical/clinical diagnosis of tuberculosis index cases without microbiological testing; C - No description of the derivation of the cohort;
[c] A*- Reported testing for and numbers of tuberculosis cases in children at baseline; B*- Reported prevalent tuberculosis as an exclusion criteria for incident tuberculosis; C - No demonstration of lack of tuberculosis disease at baseline visit
[d] A*- Prospective Cohort; B* - Adjusted odds ratio; C - Retrospective Cohort; D - Adjusted Odd Ratio not specified; E - nothing specified;
[e] A*- Microbiological testing; B* - Radiographical and clinical (must have both); C - Radiographical or clinical (only 1); D - No description;
[f] A* - Yes (at least 3 months) after exposure to infectious patient with mycobacterium tuberculosis*; B - No; C - Information not provided;
[g] A* - If prospective, all children exposed were evaluated for tuberculosis during follow-up; B* - If prospective, <= 10% of children exposed lost to follow up; C* - If retrospective, number of children lost to follow-up or excluded is reported and <=10%; D - If retrospective or prospective, greater than 10% lost to follow up; E - If prospective or retrospective, number of children lost to follow up not reported.

Table S7. Demographic characteristics of participants administered and not administered preventive treatment.

| **Characteristic** | **Not administered INH preventive treatment** | **Administered INH preventive treatment** |
|---|---|---|
| | **n, %** | **n, %** |
| Total individuals evaluated for incidence | 4,134 | 722 |
| Total person-years follow-up | 10,375 | 1,663 |
| Median follow-up time (IQR) | 2.3 (1.4-4) | 2.1 (1.4-3.3) |
| Age group, years | | |
| <10 | 515 (13) | 326 (45) |
| 10-19 | 493 (12) | 291 (40) |
| 20-29 | 794 (19) | 36 (5) |
| 30 and over | 2,294 (56) | 69 (10) |
| Sex | | |
| Male | 1,907 (47) | 339 (49) |
| Female | 2,170 (53) | 354 (51) |
| QuantiFERON or tuberculin skin tested | 1,381 (33) | 169 (23) |

Figure S1. Map of Location of Included Studies.

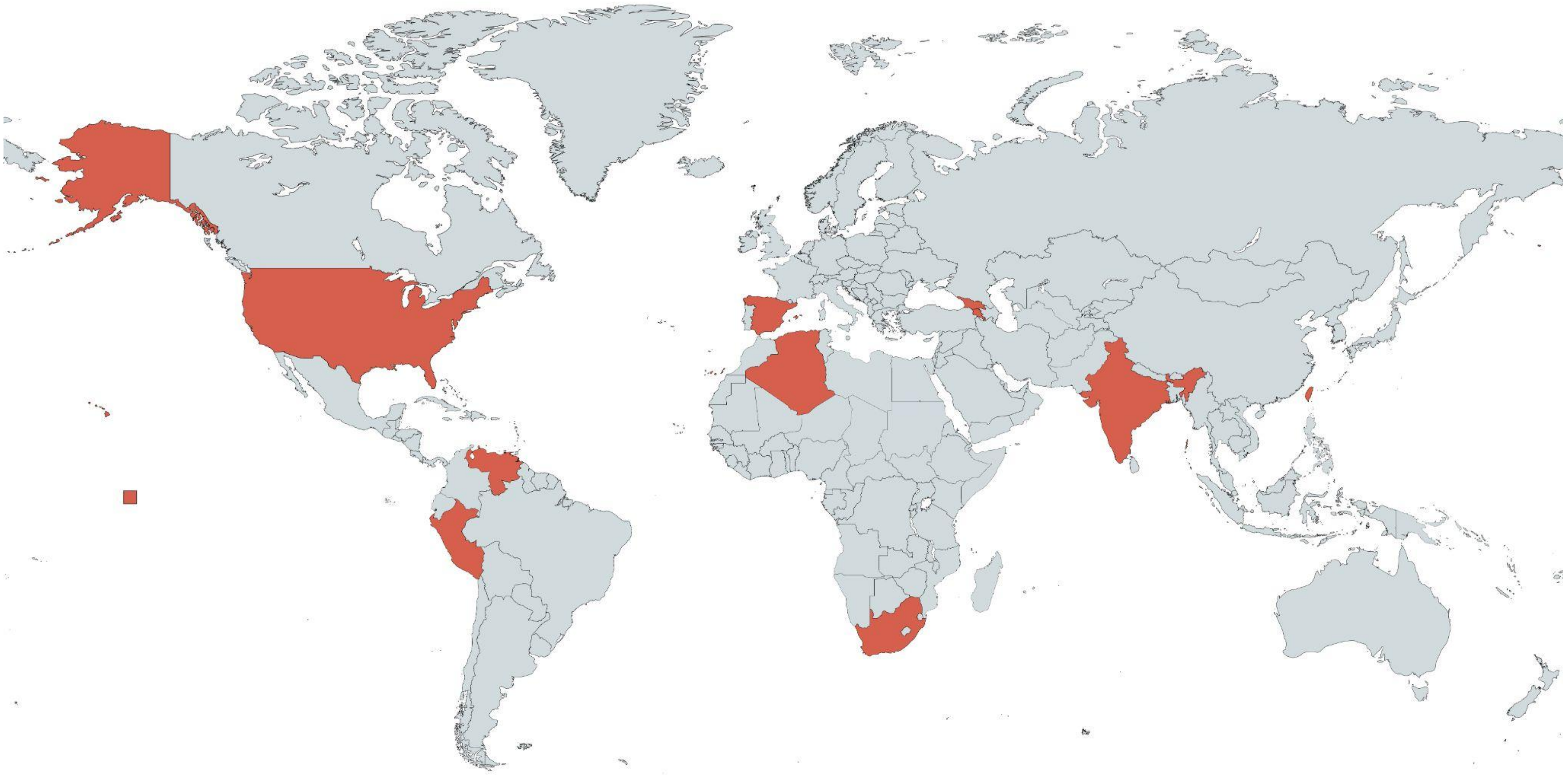

Figure S2. Two- and four-year follow-up for tuberculosis, stratified by isoniazid TPT given

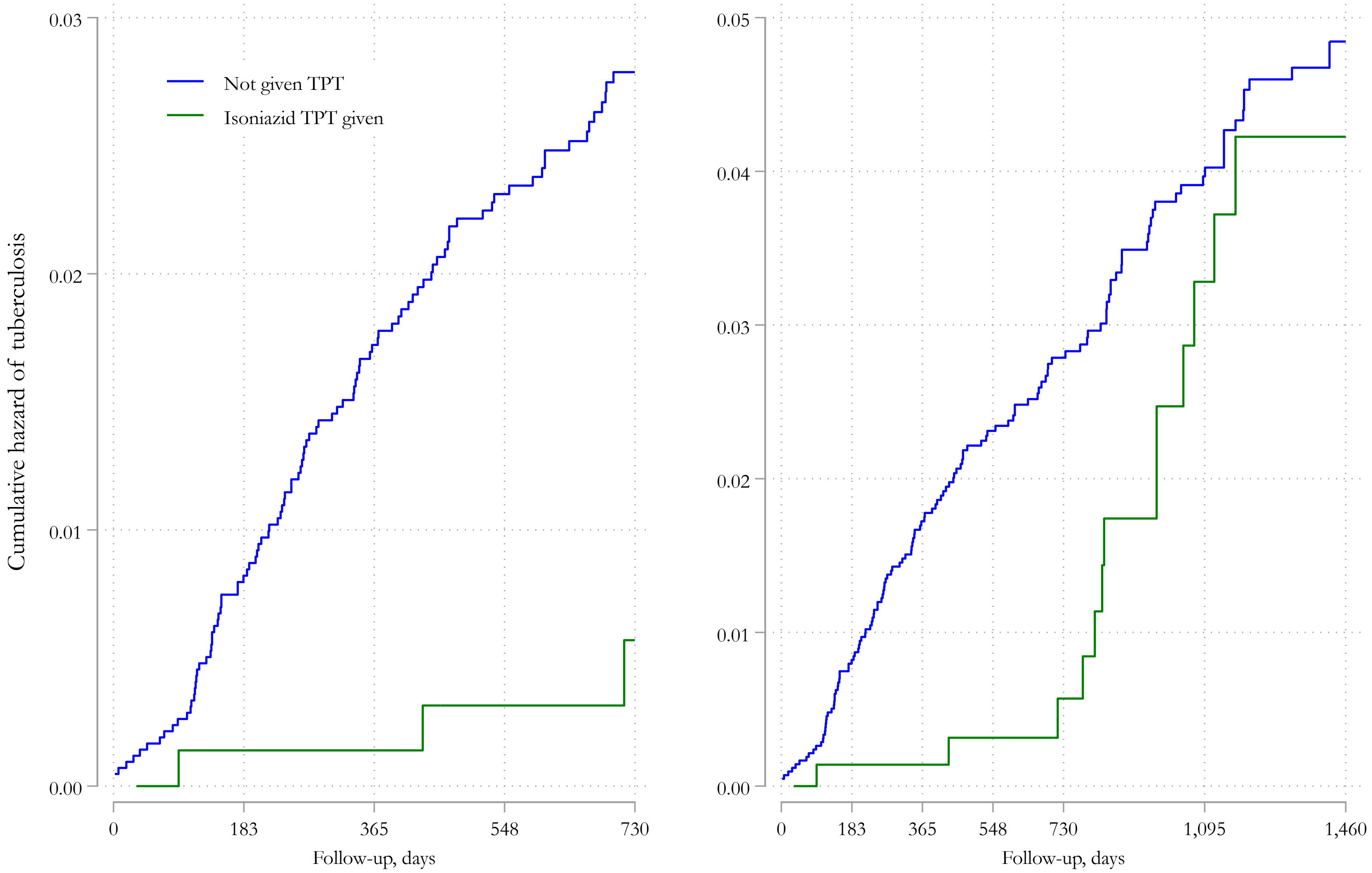


†This graph is a Kaplan Meier curve, unadjusted for study-level random effects. Therefore, as you go along the X-axis and follow-up, there are fewer and fewer patients included. This could lead to other biases, especially after 2 years of follow-up.
